# Deep Learning-Based Missing Value Imputation for Heart Failure Mortality risk Prediction Data from MIMIC-III: A Comparative Study of DAE, SAITS, and MICE+LightGBM

**DOI:** 10.64898/2026.02.10.26345979

**Authors:** Shilpa Sharma, Mandeep Kaur, Savita Gupta

## Abstract

**Background:** Electronic Health Records(EHR) are very crucial for Clinical Decision Support Systems and for proper care to be delivered to ICU heart failure patients, there is often missing data due to monitoring device errors thus the need for robust imputation methodologies.

**Objective:** To compare and evaluate three different methodologies for imputing missing data for heart failure patients from the MIMIC-III database: Denoising Autoencoder (DAE), Self-Attention Imputation for Time Series (SAITS), and Multiple Imputation by Chained Equations (MICE) with LightGBM.

**Methods:** Analysis of 14,090 ICU admissions for patients with heart failure was performed using data from the MIMIC-III database. Features were selected based off of clinical relevance, and 19 clinical features were selected through a combination of Random Forest analysis, correlation analysis, and Mutual Information. The introduction of artificial missing values of 20%, 30%, and 50% was applied to the data set, and then 3 imputation methodologies were evaluated with the DAE, SAITS, and MICE+LightGBM. The performance of each imputation methodology was evaluated using Mean Absolute Error (MAE), Root Mean Square Error (RMSE), and Normalized Root Mean Square Error (NRMSE).

**Results:** Both DAE and SAITS had superior performance on the imputation of missing values across all percentages of missing values. At 20% missingness, DAE had mean MAE = 0.004967, RMSE = 0.005217, and NRMSE = 3.260893 while SAITS had mean MAE = 0.005461, RMSE = 0.005797, and NRMSE = 3.244695; thus MICE+LightGBM resulted in a higher number of errors. At 50% missingness, the SAITS methodology demonstrated the best performance followed by DAE and MICE+LightGBM methods demonstrated decreased performance. The deep learning methodologies maintained a consistent level of accuracy between the clinical variables measured.

**Conclusions:** Our analysis indicates that deep learning-based imputation methodologies significantly outperform traditional methodologies for imputing missing values in ICU heart failure data thus supporting the implementation of these methodologies into Clinical Decision Support Systems for heart failure patients.

## 1 Introduction

Heart failure is one of the most common causes of morbidity and mortality worldwide, and ICU admissions are among the most serious events requiring ongoing physiological measurements and rapid clinical responses. The available EHR data for ICU use, which are the only longitudinal data collected continuously during all ICU visits, comprise vital signs and laboratory information (both of which are collected continuously), medications provided, and eventual clinical outcomes for each patient. Unfortunately, for various reasons (i.e., a lack of a consistent measurement schedule, equipment failure, and prioritizing clinical assessments), much of these data are often incomplete [1][2][3]. As such, when data researchers use historical data from clinical databases in predictive modelling, clinical decision support systems, or retrospective studies, they encounter significant challenges due to the prevalence of missing values in clinical time-series data [4][5]. In fact, the prevalence of missing values in MIMIC (an ICU clinical database) can reach up to 80% for certain laboratory values [6][7]. Traditional approaches to addressing missing data, such as complete-case analysis, simple mean imputation, and Last Observation Carried Forward (LOCF), can introduce significant bias, reduce statistical power, and fail to capture the complex temporal and multivariate dependencies inherent in clinical data [8][9]. Consequently, there is typically a great deal of interest in advanced approaches that leverage the rich nature of EHR data to derive accurate, clinically plausible estimates of missing data points.

In the last few years, the number of deep learning techniques available for filling in missing values in patient charts for clinical trials has increased owing to advances in technology and new research methods. For example, considering the mortality rates for patients in both the MIMIC-III and MIMIC-IV patient populations, many researchers have recently published their findings using various forms of deep learning to develop new models that incorporate either or both imputation and mortality rate-based predictions (i.e., VAEs + RNNs) [1]. Similarly, researchers have developed GANs that help resolve the issues of both imputation and balancing between classes with respect to the number of records in their datasets [2]. Over the last few years, SAITS, a transformer-based architecture, has achieved state-of-the-art performance in the imputation of clinical time-series data using MIMIC-IV monitoring datasets [9].

Although numerous successful cases exist, the literature also identifies substantial opportunities for enhancement in several areas. Many studies using deep learning have focused primarily on either the general population of ICU patients or have simply focused on one or two conditions (i.e., sepsis) [10][11]. Consequently, few studies have been conducted on patients with heart failure (which is an important population to study because they have specific pathophysiological characteristics). Additionally, few studies have compared various hybrid statistical and machine learning approaches for imputation and for analysing such patients using aggregated time-series data rather than cross-sectional or longitudinal data (e.g., gradient-boosting + MICE). Finally, there is a need for a systematic evaluation of imputation performance across a range of missing data rates and a large number of different clinical variables (i.e., vitals, lab tests, labs, and calculated statistics) to support clinical decision-making around the implementation of algorithms for either or both features of missing data.

To address the lack of information regarding imputation algorithm performance in patients with heart failure, we developed a rigorous comparative analysis of three different imputation algorithms:DAE, SAITS, and multiple imputation by chained equations MICE+LightGBM on a large heart failure cohort obtained from MIMIC-III. Our main research questions are as follows:

1. How accurate are different types of imputation in heart failure data with varying degrees of missingness (20%, 30%, and 50%)?
2. Do deep learning algorithms provide better performance than traditional statistical and machine learning methods?
3. How do the algorithms perform at both a global level and an individual variable level?
4. What evidence-based guidelines can be provided to support imputation strategy selection for use in clinical decision support systems for heart failure?
5. How does the SHAP method improve model interpretability by explaining the influence of each individual feature on the model’s predictions and offering practical insights to support clinical decision-making?

## 2 Related work

Conventional imputation methods, such as mean, median, and mode imputation, often fail to adequately account for temporal information or patient-specific variation [12][13]. Hence, regression imputation and expectation-maximization techniques are also being adapted to account for temporal dependencies and through time series regressions; however, these methods require that assumptions about the distribution of the data are met, which is not always the case in ICU data sets [14][15][16][17]. Machine learning techniques are increasingly recognised as having advantages over traditional statistical techniques, as they can accurately model nonlinear relationships and temporal dynamics [18][19]. Moreover, a variation of k-nearest neighbours imputation that incorporates temporal proximity within the dataset has been developed. In addition, tree-based and ensemble models can effectively address heterogeneous data types and complex missing-value patterns, thereby improving imputation accuracy [20][21]. Recurrent neural networks, long short-term memory networks and gated recurrent units are extremely well suited to the clinical heart failure ICU time series, as they can model the sequential dependencies in the data as well as account for the varying lengths of sequences and missing intervals, all the while providing much greater accuracy for reconstructing clinically important missing measurements [22].

Additionally, the development of advanced generative models, such as autoencoders and generative adversarial networks, to model latent temporal representations of the data will enable realistic imputation of missing segments, and combining multiple imputation approaches can yield quantitative uncertainty estimates and improve the robustness of clinical conclusions [23][24][25]. Another emerging trend is the development of hybrid approaches that combine statistical and machine learning techniques, with some incorporating expert knowledge of the underlying process (e.g., physiological constraints and clinical guidelines) to address challenges in data imputation while also ensuring alignment with contemporary clinical workflows [26][27]. Despite advances in using machine learning algorithms to address data gaps, challenges remain in handling irregular sampling, ensuring real-time imputation when data are continuously monitored, and ensuring the generalizability of the imputation model across all patient populations. Accordingly, future research should focus on the development of interpretable and clinically validated imputation models that can be incorporated into decision support systems in the ICU to ultimately improve the management of heart failure patients [28][29][30].

None of the studies mentioned above attempted to improve the interpretability of the models produced by utilising the SHAP methodology[1][2][6][8][31]. It provides a unified measure for interpreting each feature’s contribution to a model’s predictions by assigning additive importance values based on cooperative game theory. It enables transparent, consistent explanations at both global and individual levels, facilitating a deeper understanding of complex machine learning models. In this research, SHAP values are used to elucidate how clinical variables influence imputation outcomes, supporting evidence-based selection of imputation strategies in heart failure data analysis[32]. From the above, it is clear that most approaches focus on evaluating the imputation performance of ML models on medical datasets. Significantly, no studies focus on enhancing the explainability of their models, primarily using SHAP for missing-value imputation.

## 3 Materials and Methods

### 3.1 Missing Data Mechanisms in Clinical Settings

Missing data in EHRs arise from three primary mechanisms: missing completely at random (MCAR), missing at random (MAR), and missing not at random (MNAR) [31]. In ICU settings, missingness is rarely MCAR; rather, it often reflects clinical decision-making processes in which sicker patients receive more frequent monitoring (MAR) or in which specific measurements are omitted due to clinical judgment about their necessity (MNAR) [33]. Understanding these mechanisms is crucial for selecting appropriate imputation strategies, as methods that assume MCAR may introduce bias when applied to MAR or MNAR scenarios common in clinical practice [34].

### 3.2 Data Source and Cohort Selection

#### 3.2.1 MIMIC-III Database

The MIMIC-III database contains de-identified patient data from 61,532 intensive care unit (ICU) admissions to the Beth Israel Deaconess Medical Centre over a period of time (2001-2012). Unlike open databases, access to MIMIC-III requires researchers to complete research ethics training provided by the PhysioNet platform and sign a Data Use Agreement (DUA). The PhysioNet Platform is managed by members of the MIT laboratory for Computational Physiology [35]. MIMIC-III clinical database is a comprehensive repository of clinical data for use in clinical informatics research, including demographics, vital signs, laboratory test results, medications, procedures, diagnoses, and clinical outcomes [36]. The MIMIC-III clinical database is a well-validated resource widely used for research in clinical informatics.

#### 3.2.2 Heart Failure Cohort Construction

To analyse heart failure admissions, we utilised the DIAGNOSES ICD table containing the International Classification of Diseases, Ninth Revision (ICD-9) diagnosis codes for heart failure: 4280 (congestive heart failure, unspecified), 42820 (systolic heart failure, unspecified), and 42821 (acute systolic heart failure). These codes were utilised based on their clinical relevance and prior use in heart failure studies conducted using MIMIC-III [37]. Only unique hospital admissions (HADM ID) associated with these codes remained in our data.

Heart failure hospitalisations and ICU lengths of stay were evaluated using the ICUSTAYS data. ICU admission times (INTIME) were used as temporal anchors for all subsequent data extraction. A Python-based data processing environment was established to handle the large-scale MIMIC-III dataset, using the pandas library for data manipulation and NumPy for numerical operations, while chunk-based processing enabled efficient handling of massive event tables. Heart failure admissions were first identified from the DIAGNOSES ICD table using ICD-9 discharge codes. Each ICU stay was then linked to its corresponding heart failure admission through the ICUSTAYS table, with the INTIME field serving as the reference point for time alignment.

A set of 21 clinically relevant physiological features was selected based on prior literature, including vital signs extracted from the CHARTEVENTS table (e.g., heart rate, respiratory rate, blood pressure, and temperature) and laboratory measurements from the LABEVENTS table (e.g., hemoglobin, creatinine, glucose, electrolytes, and coagulation markers). ITEMIDs were mapped to human-readable feature names to ensure interpretability. Clinically plausible physiological ranges were defined using clinical judgment and published references, and values outside these limits were removed to eliminate implausible measurements. To manage data scale, CHARTEVENTS and LABEVENTS were processed in two-million-row chunks, retaining only relevant ITEMIDs, removing nonnumeric entries, and converting timestamps into datetime format. Finally, event records were temporally filtered by merging them with ICU stay information, retaining only measurements recorded within the first 24 hours of ICU admission and for patients older than 16 years to maintain temporal consistency and prevent data leakage. After applying the data pre-processing steps and data quality criteria, 14090 ICU stays in the final cohort of study patients were analysed. The complete methodology of this study is shown in Figure 1.

**Fig. 1:**
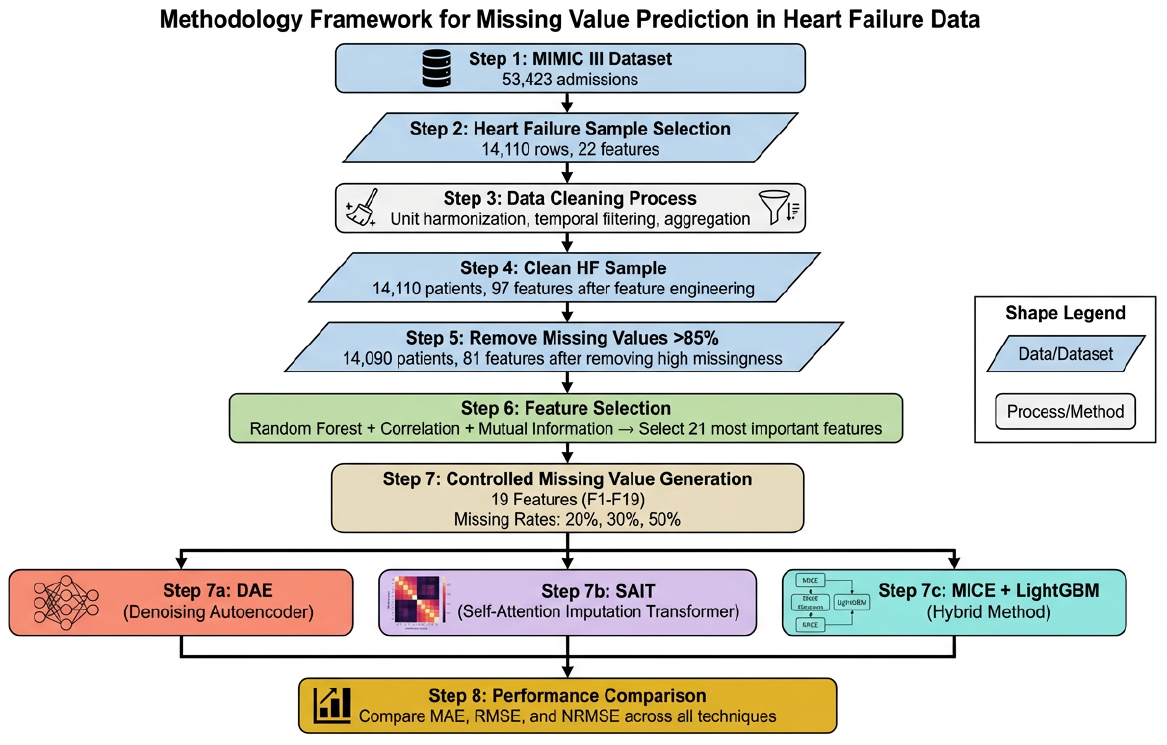
Methodology

### 3.3 Data Preprocessing Pipeline

To ensure the integrity (quality, temporal validity, and clinical relevance) of our data, we implemented a rigorous 6-step pre-processing pipeline. The following are the first steps:

1. Unit Harmonisation – Temperature measurements in Fahrenheit were converted to Celsius. Following unit harmonisation, physiological range filters were applied to remove remaining outliers and implausible values.
2. Time-Series Aggregation – For each ICU stay and variable, measurements from the first 24 hours were summarised using four statistical descriptors: mean, minimum, maximum, and standard deviation. This transformed irregular time series into fixed-length feature vectors suitable for machine learning.
3. Reshaping Data Set - Pivot operations were performed to change the long format into a wide format by converting it into a pivot table. Each row represents a unique ICU stay, and each column has one aggregated feature. This produced a statistical dataset with 14,110 rows and 97 features.
4. Missing Value Filtering – All features with over 85% of their values missing were removed. The Hospital Expired Flag (HOSPITAL EXPIRE FLAG) was included in the ADMISSIONS table. The number of records in the dataset is 14,090 ICU Stays and 81 total features (78 clinical and 3 demographic). The complete list of 81 features is presented in Table 1.
5. Feature Selection – The features that are to be used as predictors are 19 clinical and 2 demographic, selected from 81 total features using a combination of three methods: (1) the importance of each feature variable using Random Forest with classification accuracy; (2) Pearson correlation coefficients to eliminate redundant features; and (3) mutual information scores to determine which features will be most predictive of outcome as shown in Figures 2–4 and 19 features are used as target variable in this research are shown in a Table 2.
6. Final Dataset Generation – The final preprocessed dataset comprised 14,090 heart failure ICU stays and 81 aggregated features, exported as a CSV file for imputation experiments.

**Table 1:**
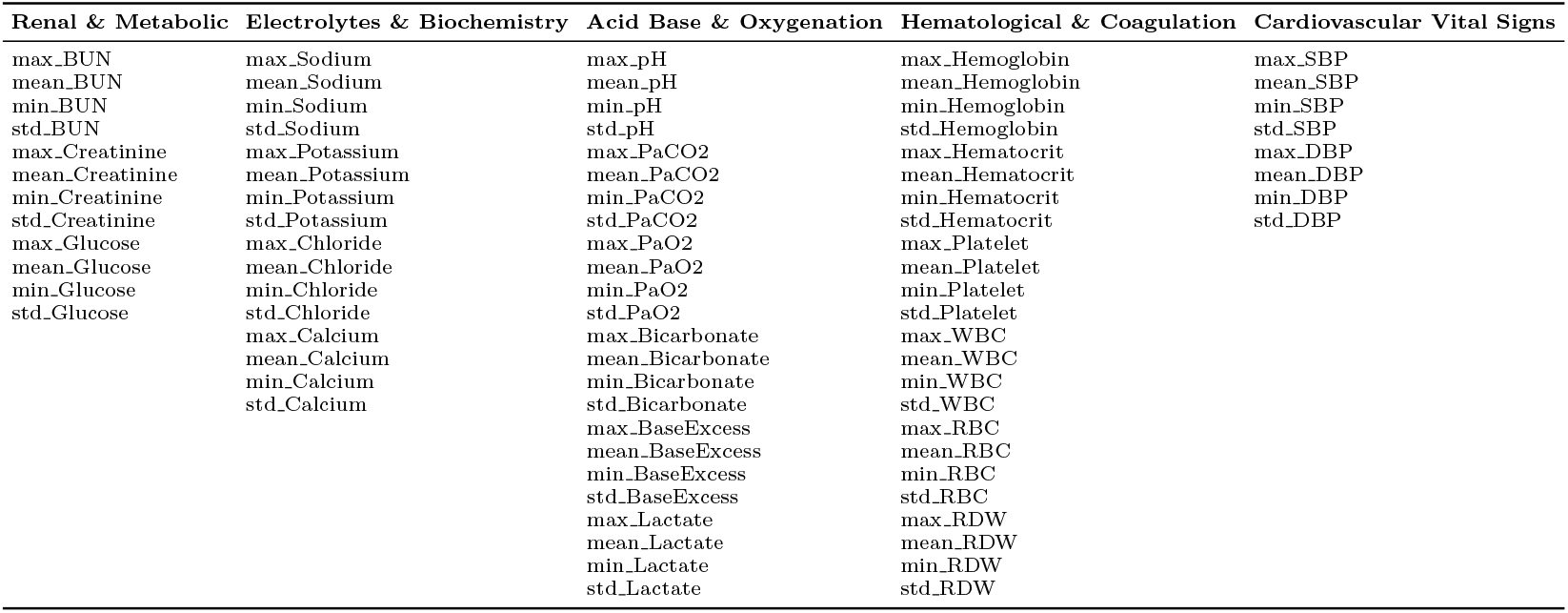
List of 81 predictor variables grouped by physiological category.

**Table 2:** 19 Target variables Categories and Value Ranges.

| Category | Features | Values Range |
| --- | --- | --- |
| Lactate measurements | Mean_Lactate, max_Lactate, min_Lactate, std_Lactate | 0.1 to 30 |
| Acid-base balance | Mean_pH, max_pH, min_pH, std_pH | 6.8 to 7.8 |
| Renal function | Max_BUN, min_BUN | 1 to 200 |
| Electrolytes | Mean_Bicarbonate, min_Bicarbonate, max_Bicarbonate | 5 to 50 |
| Hematology | Mean_RDW, max_RDW, min_RDW, mean_Platelet | RDW (10 to 30), Platelet (5 to 1000) |
| Respiratory | Mean_Paco2, std_Pao2 | Paco2 (10 to 120), Pao2 (20 to 600) |

**Fig. 2:**
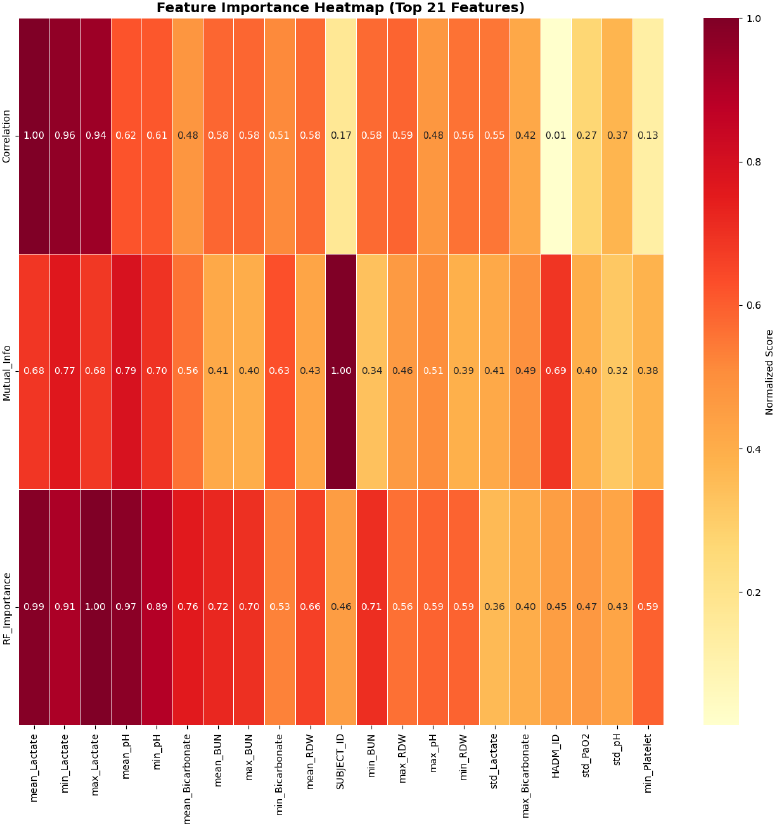
Feature importance heatmap of 21 features

**Fig. 3:**
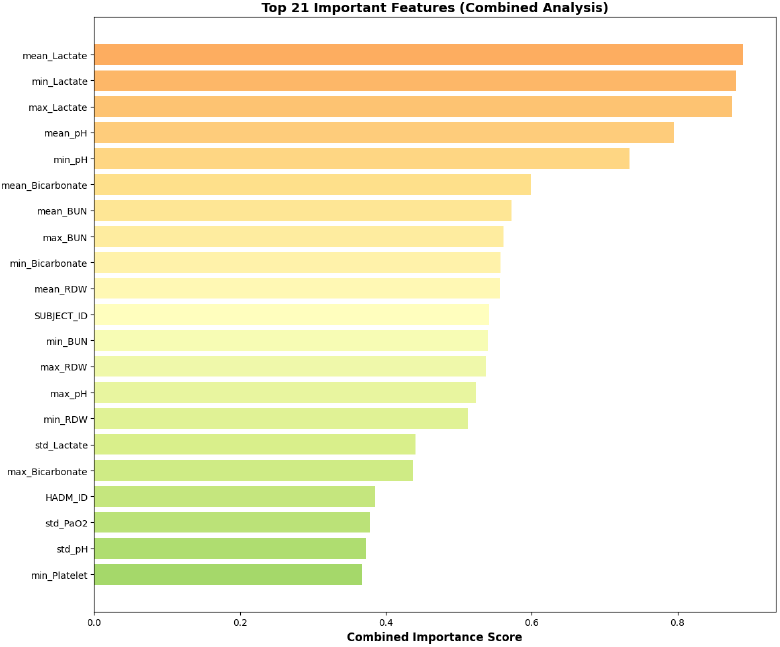
combined analysis: Random Forest, Mutual Information score and Pearson correlation score for 21 important features selection.

**Fig. 4:**
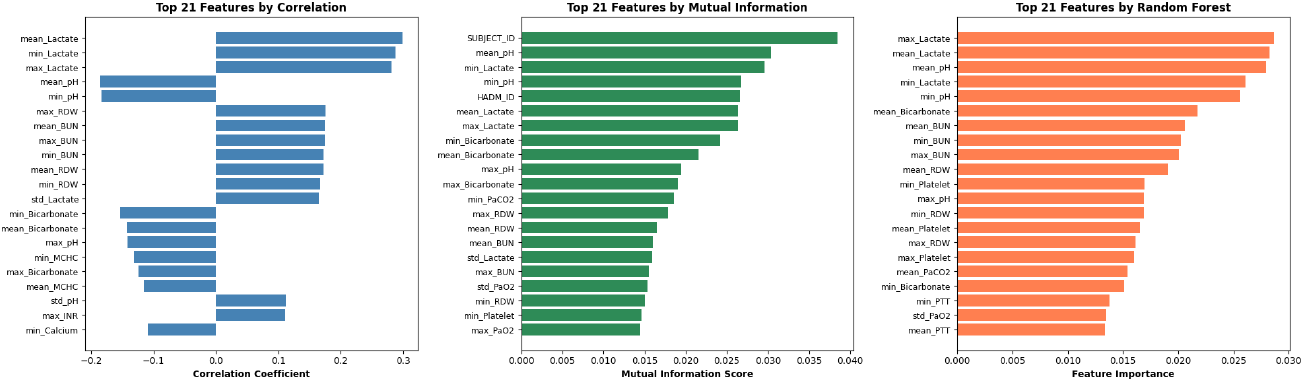
leftone represents the feature selected by correlation coefficients, middleone represents the feature selected by Mutual information, and rightone represents the feature selected by Random Forest.

Table 1 presents the 19 target variables with their values, in this study. These features capture key pathophysiological processes in heart failure including tissue perfusion (lactate), acid-base disturbances (pH, bicarbonate, PaCO2), renal dysfunction (BUN), and hematological abnormalities (RDW, platelets).

### 3.4 Missing Value Introduction Protocol

To determine how well imputation performs under controlled conditions, we created missing data for each of the 19 target variables by selecting three different levels of missingness (20%, 30%, and 50%). This procedure is often used in imputation benchmarking research [9][38] to quantitatively compare the imputed and true (or ground truth) values. Missing values were created using the missing completely at random (MCAR) method to provide an unbiased examination of the statistical performance of the imputation algorithms. At each of the three missingness levels, values from each feature were randomly selected to be masked, ensuring that they were masked based on their original proportion. This resulted in the production of three experimental datasets, with the true values of all masked samples known, allowing for the exact computation of imputation errors.

### 3.5 Imputation Algorithms

#### 3.5.1 Denoising Autoencoder (DAE)

The DAE has an encoder-decoder architecture with a fully connected layer. The encoder compresses the input feature space to a low-dimensional latent representation, and the decoder recreates the input feature space from the low-dimensional latent representation. The DAE is trained to minimize the reconstruction error in the observed data; therefore, denoised inputs are obtained by recovering the missing values. Training employed mean squared error (MSE) loss on observed values only, with missing values masked during the loss calculation. The Adam optimizer was used with a learning rate scheduling. Dropout regularization was applied to prevent overfitting. The trained model was then used to impute the missing values in the test datasets.

#### 3.5.2 Self-Attention Imputation for Time Series (SAITS)

The SAIT reconstructs missing values in time-series data by learning temporal relationships among observed measurements. In Figure 5, the input sequence contains observed values at (t 1, t 2, t 3) and (t 6), while the values at (t 4) and (t 5) are missing. A binary mask vector is first created to indicate which positions are observed (1) and which are missing (0). Because neural networks cannot process blanks, the missing entries are temporarily replaced with placeholder values (such as zeros or mean values), and positional encoding is added to preserve the time order of the sequence. The resulting sequence is passed through a Transformer encoder, where the self-attention mechanism allows each time step to learn from all other time steps in the sequence. Through this process, the missing positions attend to the observed values and capture temporal trends and contextual dependencies. The encoder produces contextual representations that are fed into a prediction layer, which estimates the values at the masked positions. During training, the model minimizes reconstruction error only at the missing locations, enabling it to learn accurate imputation patterns. Finally, the predicted values replace the missing entries, producing a complete time series in which only the original missing positions (t 4) and (t 5) are filled while the sequence structure remains unchanged.

**Fig. 5:**
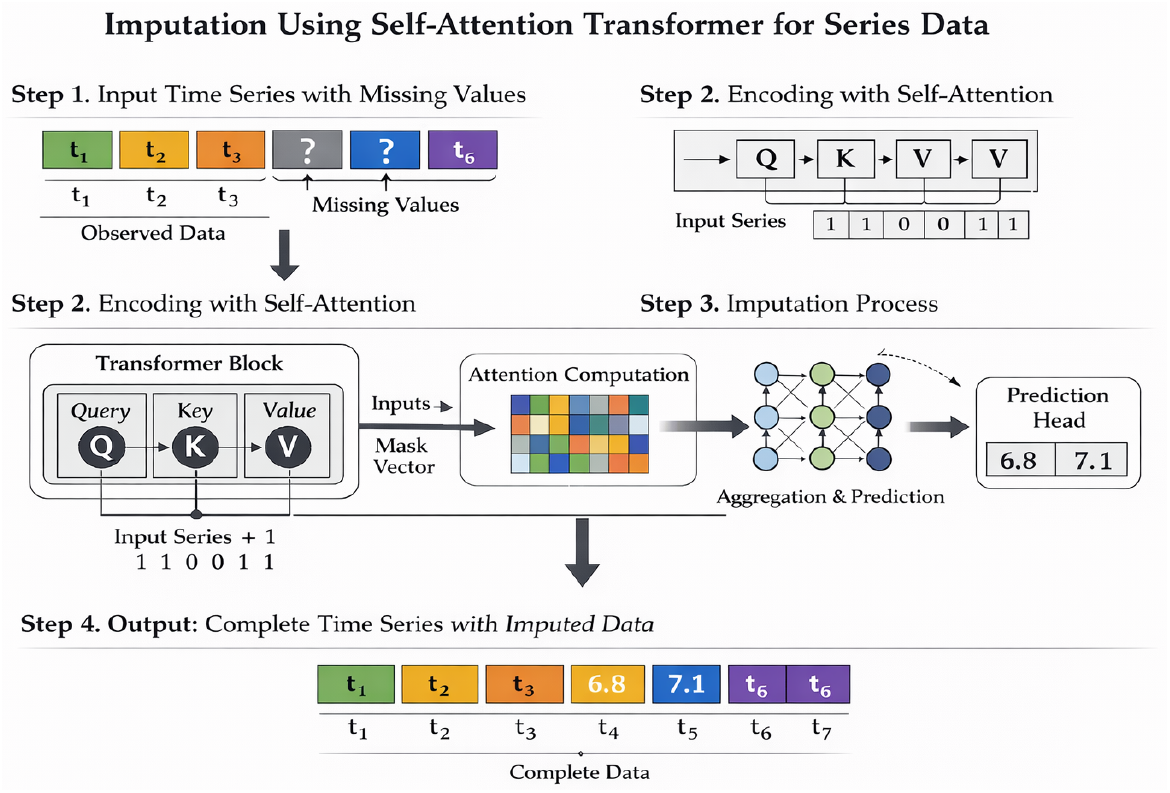
Imputation Working Diagram of SAIT

#### 3.5.3 Multiple Imputation by Chained Equations with LightGBM (MICE+LightGBM)

MICE+LightGBM combined the principled multiple imputation framework with gradient boosting regression. For each variable with missing values, a LightGBM regression model was trained using all other variables as predictors. This process was iterated cyclically through all variables with missing values until convergence. Light-GBM hyperparameters were tuned for each variable, including the number of leaves, the learning rate, and the number of boosting rounds. Multiple imputation generated five complete datasets, and the results were pooled to obtain the final imputed values. This approach combined MICE’s uncertainty quantification with LightGBM’s strong performance on tabular data.

### 3.6 Evaluation Metrics

The imputation quality is typically assessed using metrics that quantify the discrepancies between the imputed and true values. Mean Absolute Error (MAE) is an average of absolute differences and is an interpretable metric related to scale. Because of the squaring of errors, the Root Mean Square Error (RMSE) places a greater penalizing weight on larger errors, making the RMSE sensitive to outliers. The Normalized Root Mean Square Error (NRMSE) is the RMSE divided by either the variable range or standard deviation, allowing for comparisons between feature sets measured on different scales [25]. The global NRMSE will take RMSE for all masked values and will normalize it to the standard deviation of the actual observation. For imputation evaluation in clinical settings, assessments should take into account both the statistical accuracy of the imputed value as well as the clinical plausibility of the imputed value. Imputed values will have a lower MAE and lower RMSE, indicating a better statistical fit; however, physiologically implausible imputed values will produce insufficient clinical meaning between the imputed value and other variable value relationships [26]. Additionally, downstream task performance (e.g., mortality prediction accuracy) provides an important complementary evaluation of the imputation quality [27].

Imputation performance was assessed using three complementary metrics:

#### Mean Absolute Error (MAE)

The mean absolute error is defined as:

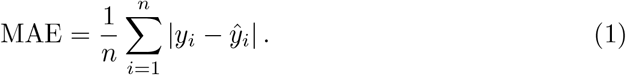

where *y*_*i*_ is the true value and observation. *ŷ*_*i*_ is the imputed value for the *i*-th missing

#### Root Mean Square Error (RMSE)

The root-mean-square error (RMSE) is defined as

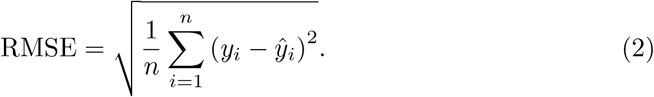

#### Global Normalised Root Mean Squared Error (NRMSE)

The global normalized root mean squared error (NRMSE) is defined as

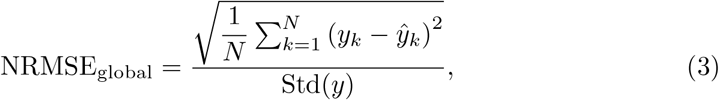

where *y*_*k*_ are the true masked values, *ŷ*_*k*_ are the imputed values, *N* is the number of masked samples, and Std(*y*) denotes the standard deviation of the true masked values.

Mean MAE (feature-wise average):

The mean MAE over all features is defined as:

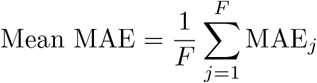

where, for each feature *j*,

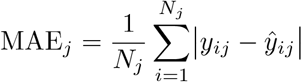

with:

- *F* : number of features,
- *j*: feature index (*j* = 1, …, *F*),
- *N*_*j*_: number of masked samples for feature *j*,
- *y*_*ij*_: true value of feature *j* for sample *i*,
- *ŷ*_*ij*_: imputed (predicted) value of feature *j* for sample *i*.

Mean RMSE (feature-wise average) — Formula Each feature’s RMSE is computed independently, and then the RMSE values are averaged across features.

For each feature *j* ∈ {1, …, *F*}, the root mean squared error (RMSE) is defined as

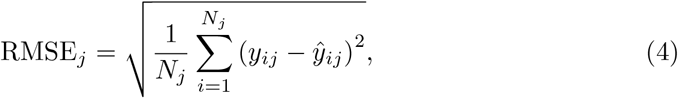

where *F* denotes the total number of features, *j* is the feature index, *N*_*j*_ is the number of masked samples for feature *j, y*_*ij*_ is the true value of feature *j* for sample *i*, and *ŷ*_*ij*_ is the corresponding imputed value. Metrics were calculated both globally (averaged across all features and observations) and feature-specifically (for each of the 19 selected variables). This dual-level evaluation provided a comprehensive assessment of imputation quality across diverse physiological measurements.

### 3.7 Experimental Design

For each missingness level (20%, 30%, 50%), the following procedure was executed:

1. Randomly mask the specified percentage of values across 19 target variables
2. Apply each imputation algorithm (DAE, SAITS, MICE+LightGBM) to the masked dataset
3. Calculate MAE, RMSE, and NRMSE by comparing imputed values to ground truth
4. Compute global metrics (averaged across all features) and feature-specific metrics
5. Repeat the process with different random seeds to ensure reproducibility
6. Applied SHAP (SHapley Additive exPlanations) to the SAITS imputation model to quantify and rank feature contributions to missing value reconstruction, enabling verification that the imputation process preserved clinically meaningful physiological relationships within the heart failure ICU dataset. Experiments were performed on a computing cluster consisting of NVIDIA GPUs to explore deep learning models. Results were not tested for statistical significance; however, because of the very large sample size (14,090 ICU), combined with the control in our experiment, there was adequate statistical power to detect significant differences.

## 4 Results

### 4.1 Global Performance Comparison

Table 3 presents the global imputation performance across all three methods and missingness levels. Global metrics were computed by averaging MAE, RMSE, and NRMSE across all 19 features.

**Table 3:** Global RMSE and NRMSE for DAE, SAIT and MICE+LightGBM models and masking percentages.

| Model | Masking (%) | Global RMSE | Global NRMSE |
| --- | --- | --- | --- |
| DAE | 20 | 0.006331 | 3.260893 |
| SAIT | 20 | 0.006299 | 3.244645 |
| MICE+LightGBM | 20 | 0.221432 | 0.222056 |
| DAE | 30 | 0.006327 | 3.263749 |
| SAIT | 30 | 0.006613 | 3.403345 |
| MICE+LightGBM | 30 | 0.265134 | 0.266477 |
| DAE | 50 | 0.006340 | 3.278050 |
| SAIT | 50 | 0.005076 | 2.618904 |
| MICE+LightGBM | 50 | 0.376995 | 0.377731 |

At 20% missingness, there was virtually no difference in performance between DAE and SAITS. The RMSE values were nearly identical (0.006331 versus 0.006299), and the NRMSE values were also nearly identical (3.260893 versus 3.244645). However, as the missingness increased to 30%, both techniques continued to perform stably, with a small but apparent difference in performance. The DAE displayed minimal degradation (the RMSE decreased from 0.006331 to 0.006327), whereas the NRMSE increased from 3.260893 to 3.2637349. Likewise, the performance of SAITS was virtually unchanged; however, the NRMSE increased slightly from 3.244645 to 3.403345. However, the absolute differences between the two techniques’ performance were almost undetectable. When the missingness rate increased to 50%, SAITS outperformed DAE. The DAE had an RMSE of 0.00634 and an NRMSE of 3.27805. DAE performed comparably to SAITS with respect to NRMSE (2.618904 for SAITS and 3.27805 for DAE). Thus, although there are significant absolute differences in performance between the methods, the methods exhibit comparable levels of normalised performance.

### 4.2 Feature-Specific Performance Analysis

The model performance across all features with 20% missing data is presented in Table 4. At 20% missingness, DAE performed consistently better than MICE+LightGBM over all features. For lactate measurements, DAE produced an MAE of 0.002421 for mean_Lactate, while MICE+LightGBM hadan MAE of 0.083502 improvement. Using the same methods, the MAEs for the pH variables from DAE and MICE+LightGBM were similar; mean_pH was therefore calculated to have an MAE of 0.00746 for DAE and an MAE of 0.148502 improvement for MICE+LightGBM. SAITS demonstrated competitive performance with DAE, with some features showing superior SAITS performance (e.g., min_Lactate: SAITS MAE=0.001281 vs. DAE MAE=0.002637) and others favoring DAE (for example, mean_pH: DAE MAE=0.00746 vs. SAITS MAE=0.01069). The variability in the relative performance across features suggests that both deep learning methods capture different aspects of the underlying data distribution. MICE+LightGBM showed substantially higher MAE across all features, with errors ranging from 0.023507 (max_BUN) to 0.296122 (std_pH). The particularly poor performance on standard deviation features (std_Lactate, std_pH, std_PaO2) suggests that the hybrid statistical-machine learning approach struggled to capture the distributional properties of temporal variability measures.

**Table 4:** Feature-Specific Imputation Performance with 20% Missing Values.

| Feature | Mask (%) | DAE |  |  | MICE + LightGBM |  |  | SAITS |  |  |
| --- | --- | --- | --- | --- | --- | --- | --- | --- | --- | --- |
|  |  | MAE | RMSE | NRMSE | MAE | RMSE | NRMSE | MAE | RMSE | NRMSE |
| GLOBAL | 20 | – | 0.006331 | 3.260893 | – | 0.221432 | 0.222056 | – | 0.006299 | 3.244645 |
| mean_Lactate | 20 | 0.002421 | 0.002541 | – | 0.083502 | 0.189975 | – | 0.003036 | 0.003432 | – |
| max_Lactate | 20 | 0.014850 | 0.014994 | – | 0.107838 | 0.237003 | – | 0.005911 | 0.006316 | – |
| mean_pH | 20 | 0.007460 | 0.007621 | – | 0.148502 | 0.255561 | – | 0.010690 | 0.010934 | – |
| min_pH | 20 | 0.002522 | 0.002742 | – | 0.153618 | 0.252639 | – | 0.007800 | 0.008020 | – |
| max_BUN | 20 | 0.001899 | 0.002393 | – | 0.023507 | 0.046213 | – | 0.005660 | 0.005880 | – |
| mean_Bicarbonate | 20 | 0.000612 | 0.000773 | – | 0.053989 | 0.149253 | – | 0.003837 | 0.004184 | – |
| min_Lactate | 20 | 0.002637 | 0.002843 | – | 0.134101 | 0.327357 | – | 0.001281 | 0.001649 | – |
| mean_RDW | 20 | 0.005841 | 0.006092 | – | 0.052827 | 0.229336 | – | 0.001495 | 0.001775 | – |
| mean_PaCO<sub>2</sub> | 20 | 0.002813 | 0.002933 | – | 0.066878 | 0.120071 | – | 0.006366 | 0.006843 | – |
| mean_Platelet | 20 | 0.008651 | 0.009403 | – | 0.023741 | 0.052142 | – | 0.005100 | 0.005702 | – |
| min_Bicarbonate | 20 | 0.005415 | 0.005605 | – | 0.055555 | 0.145794 | – | 0.003351 | 0.003593 | – |
| min_BUN | 20 | 0.003391 | 0.003615 | – | 0.022247 | 0.052191 | – | 0.005158 | 0.005321 | – |
| max_RDW | 20 | 0.002165 | 0.002592 | – | 0.054453 | 0.226132 | – | 0.006333 | 0.006508 | – |
| max_pH | 20 | 0.001597 | 0.001753 | – | 0.194797 | 0.318000 | – | 0.013443 | 0.013676 | – |
| min_RDW | 20 | 0.011895 | 0.012086 | – | 0.053295 | 0.234597 | – | 0.007275 | 0.007609 | – |
| std_Lactate | 20 | 0.009511 | 0.009587 | – | 0.234315 | 0.470331 | – | 0.006873 | 0.007269 | – |
| max_Bicarbonate | 20 | 0.006604 | 0.006762 | – | 0.066112 | 0.168220 | – | 0.001219 | 0.001491 | – |
| std_PaO<sub>2</sub> | 20 | 0.001364 | 0.001708 | – | 0.197191 | 0.311161 | – | 0.002193 | 0.002783 | – |
| std_pH | 20 | 0.002730 | 0.003047 | – | 0.296122 | 0.465888 | – | 0.006748 | 0.007164 | – |

#### 4.2.1 Performance at 30% Missingness

Table 5 presents the feature-wise performance of all three models at 30% missingness. At 30% missingness, the performance gap between deep learning methods and MICE+LightGBM widened. MICE+LightGBM MAE increased substantially for most features (e.g., mean_Lactate: 0.121656 at 30% vs. 0.083502 at 20%), indicating degraded performance with higher missingness. In contrast, DAE and SAITS maintained stable or slightly improved performance for many features. The DAE continued to perform well with respect to lactate measurement (mean_Lactate MAE=0.002292; max_Lactate MAE=0.014996) and pH variables (mean_pH MAE=0.007314).SAITS demonstrated significant strength for RDW measurements (mean_RDW MAE=0.001305; max)RDW MAE=0.001228; min_RDW MAE=0.001373), indicating that these machines can effectively capture patterns in hematological variables. The performance difference between the methods was greatest for the standard deviation features. For instance, std_Lactate, MICE + LightGBM achieved an MAE of 0.3127, whereas DAE achieved 0.002885, and SAITS achieved 0.00528. This resulted in improvements of 108 and 59 times for the deep learning methods, respectively.

**Table 5:** Feature-Specific Imputation Performance at 30% Missing Values.

| Feature | Mask (%) | DAE |  |  | MICE + LightGBM |  |  | SAITS |  |  |
| --- | --- | --- | --- | --- | --- | --- | --- | --- | --- | --- |
|  |  | MAE | RMSE | NRMSE | MAE | RMSE | NRMSE | MAE | RMSE | NRMSE |
| GLOBAL | 30 | – | 0.006327 | 3.263749 | – | 0.265134 | 0.266477 | – | 0.006613 | 3.403345 |
| mean_Lactate | 30 | 0.002292 | 0.002444 | – | 0.121656 | 0.293513 | – | 0.001969 | 0.002501 | – |
| max_Lactate | 30 | 0.014996 | 0.015142 | – | 0.143338 | 0.334642 | – | 0.018470 | 0.018638 | – |
| mean_pH | 30 | 0.007314 | 0.007511 | – | 0.171805 | 0.290854 | – | 0.006929 | 0.007186 | – |
| min_pH | 30 | 0.002482 | 0.002768 | – | 0.182457 | 0.288874 | – | 0.004420 | 0.004799 | – |
| max_BUN | 30 | 0.002062 | 0.002591 | – | 0.024763 | 0.050286 | – | 0.003460 | 0.003800 | – |
| mean_Bicarbonate | 30 | 0.000645 | 0.000811 | – | 0.085805 | 0.225109 | – | 0.011589 | 0.011668 | – |
| min_Lactate | 30 | 0.002453 | 0.002718 | – | 0.173981 | 0.405327 | – | 0.001788 | 0.002288 | – |
| mean_RDW | 30 | 0.005693 | 0.005999 | – | 0.083631 | 0.279189 | – | 0.001305 | 0.001591 | – |
| mean_PaCO<sub>2</sub> | 30 | 0.002986 | 0.003118 | – | 0.070211 | 0.123709 | – | 0.005149 | 0.005492 | – |
| mean_Platelet | 30 | 0.008416 | 0.009190 | – | 0.023579 | 0.047070 | – | 0.003700 | 0.004511 | – |
| min_Bicarbonate | 30 | 0.005365 | 0.005589 | – | 0.085748 | 0.220574 | – | 0.003238 | 0.004059 | – |
| min_BUN | 30 | 0.003439 | 0.003674 | – | 0.022732 | 0.053827 | – | 0.001690 | 0.002099 | – |
| max_RDW | 30 | 0.002182 | 0.002665 | – | 0.089610 | 0.282622 | – | 0.001228 | 0.001536 | – |
| max_pH | 30 | 0.001685 | 0.001844 | – | 0.231259 | 0.368559 | – | 0.005293 | 0.005660 | – |
| min_RDW | 30 | 0.011874 | 0.012081 | – | 0.082371 | 0.282202 | – | 0.001373 | 0.001648 | – |
| std_Lactate | 30 | 0.009723 | 0.009785 | – | 0.275325 | 0.551577 | – | 0.014322 | 0.014669 | – |
| max_Bicarbonate | 30 | 0.006534 | 0.006727 | – | 0.100077 | 0.237794 | – | 0.006581 | 0.006814 | – |
| std_PaO<sub>2</sub> | 30 | 0.001405 | 0.001748 | – | 0.204424 | 0.313735 | – | 0.008782 | 0.009318 | – |
| std_pH | 30 | 0.002885 | 0.003281 | – | 0.312527 | 0.443996 | – | 0.005528 | 0.006018 | – |

#### 4.2.2 Performance at 50% Missingness

All three models achieved feature-wise performance with 50% missing data, MICE + LightGBM performance dropped significantly at 50% missing with multiple features (max_pH: 0.307232, std_Lactate: 0.379076, std_pH: 0.375071), showing an MAE *>* 0.3, indicating poor performance in capturing temporal variability patterns when 50% of the data are missing. Relative to these findings, DAE and SAITS had stable performances with DAE MAE values for all features below 0.012; specifically, DAE achieved an outstanding performance on bicarbonate and BUN measurements (mean_Bicarbonate: 0.000705, min_Bicarbonate: 0.005337, max_Bicarbonate: 0.006453, max_BUN: 0.002397, min_BUN: 0.003556).SAITS’s performance above that of DAE for several features at 50% missing included an MAE of 0.001152 for min_Bicarbonate compared to 0.005337 for DAE, representing an improvement of 4.6 times. The MAE for mean_PaCO2 was 0.001331 for SAITS, while DAE had 0.003309, representing an improvement of 2.5 times. Overall, these results suggest that the SAITS attention mechanism has advantages in severely missing data situations owing to its ability to take advantage of cross-variable relationships.

### 4.3 Comparative Analysis Across Missingness Levels

Figure 9 displays the mean MAE, mean RMSE, and global NRMSE trajectories across the missing values, for DAE, SAIT and MICE+LightGBM model. The ongoing degradation of the DAE sample in terms of RMSE from (20%) 0.004967 to (30%) 0.00497 to (50%) 0.004992 demonstrated a similar trend to that of the SAITS sample’s ongoing degradation in terms of RMSE from (20%) 0.005461 to (30%) 0.005622 to (50%) 0.004538, with the near-parallel layouts of the two methods showing comparable robustness with respect to increased missingness levels. However, MICE+LightGBM exhibited a much more pronounced performance decline, and feature-specific increases in MAE underscored this point. For example, the mean MAE for MICE+LightGBM increased from approximately 0.1051 (20%) to 0.1257 (30%) to 0.1947 (50%), representing an approximate increase of 12% from (20%) to (30%) and an approximate increase of 18% from (20%) to (50%). In comparison, DAE and SAITS exhibited either stable or slightly decreasing mean MAE, indicating greater robustness than the MICE+LightGBM method. Table 7 presents the mean MAE and mean RMSE trajectories across missing values, for each model.

**Table 6:** Feature-Specific Imputation Performance at 50% Missing Values.

| Feature | Mask (%) | DAE |  |  | MICE + LightGBM |  |  | SAITS |  |  |
| --- | --- | --- | --- | --- | --- | --- | --- | --- | --- | --- |
|  |  | MAE | RMSE | NRMSE | MAE | RMSE | NRMSE | MAE | RMSE | NRMSE |
| **GLOBAL** | 50 | — | 0.006340 | 3.278050 | — | 0.376995 | 0.377731 | — | 0.005076 | 2.618904 |
| mean_Lactate | 50 | 0.002133 | 0.002315 | — | 0.210040 | 0.445842 | — | 0.004313 | 0.004530 | — |
| max_Lactate | 50 | 0.015444 | 0.015607 | — | 0.227336 | 0.444454 | — | 0.008011 | 0.008599 | — |
| mean_pH | 50 | 0.007155 | 0.007378 | — | 0.250920 | 0.407996 | — | 0.003763 | 0.004217 | — |
| min_pH | 50 | 0.002266 | 0.002647 | — | 0.243729 | 0.377878 | — | 0.006023 | 0.006262 | — |
| max_BUN | 50 | 0.002397 | 0.002977 | — | 0.027105 | 0.060688 | — | 0.001845 | 0.002268 | — |
| mean_Bicarbonate | 50 | 0.000705 | 0.000882 | — | 0.178330 | 0.358228 | — | 0.001607 | 0.001943 | — |
| min_Lactate | 50 | 0.002014 | 0.002350 | — | 0.278712 | 0.579977 | — | 0.008463 | 0.008768 | — |
| mean_RDW | 50 | 0.005472 | 0.005848 | — | 0.187403 | 0.446646 | — | 0.006484 | 0.006633 | — |
| mean_PaCO<sub>2</sub> | 50 | 0.003309 | 0.003436 | — | 0.070799 | 0.124166 | — | 0.001331 | 0.001762 | — |
| mean_Platelet | 50 | 0.008081 | 0.008945 | — | 0.024623 | 0.050195 | — | 0.003311 | 0.004499 | — |
| min_Bicarbonate | 50 | 0.005337 | 0.005600 | — | 0.175601 | 0.349803 | — | 0.001152 | 0.001463 | — |
| min_BUN | 50 | 0.003556 | 0.003805 | — | 0.024115 | 0.056000 | — | 0.004680 | 0.004898 | — |
| max_RDW | 50 | 0.002230 | 0.002754 | — | 0.197269 | 0.445033 | — | 0.002230 | 0.002660 | — |
| max_pH | 50 | 0.001758 | 0.001920 | — | 0.307232 | 0.480507 | — | 0.004345 | 0.004757 | — |
| min_RDW | 50 | 0.011689 | 0.011931 | — | 0.186577 | 0.449826 | — | 0.002267 | 0.002684 | — |
| std_Lactate | 50 | 0.009896 | 0.009958 | — | 0.379076 | 0.647950 | — | 0.006545 | 0.007082 | — |
| max_Bicarbonate | 50 | 0.006453 | 0.006687 | — | 0.201842 | 0.382942 | — | 0.005179 | 0.005560 | — |
| std_PaO<sub>2</sub> | 50 | 0.001454 | 0.001861 | — | 0.202728 | 0.306890 | — | 0.003004 | 0.003713 | — |
| std_pH | 50 | 0.003495 | 0.003929 | — | 0.375071 | 0.563159 | — | 0.011658 | 0.011828 | — |

**Table 7:**
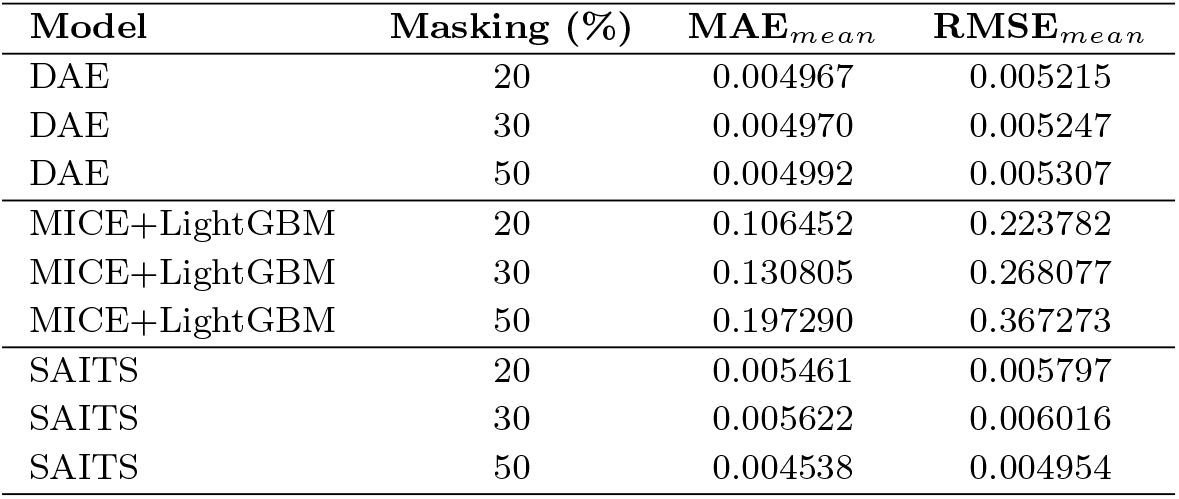
Mean MAE and RMSE across different masking percentages for each model.

### 4.4 Feature Category Analysis

Figure 6–8 represents MAE, RMSE, Global NRMSE values of DAE, SAIT and MICE+LightGBM

**Fig. 6:**
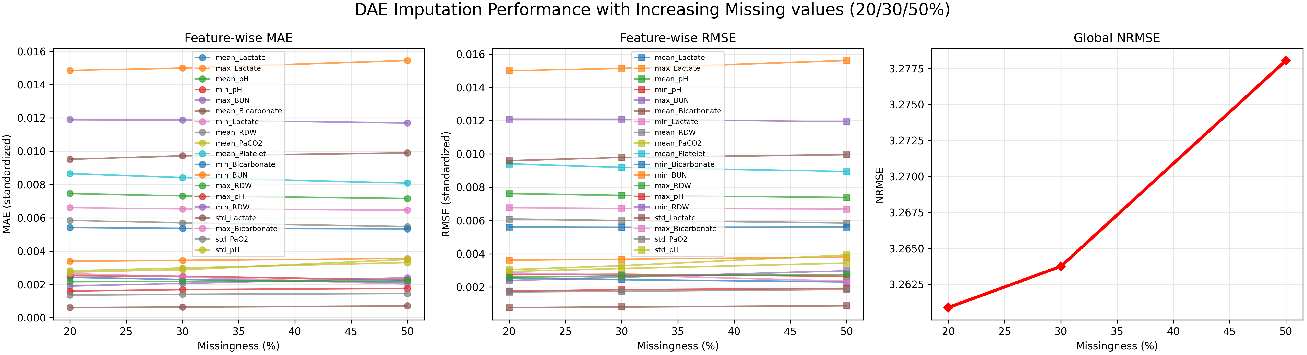
DAE Imputation Performance with Increasing Missing Values

**Fig. 7:**
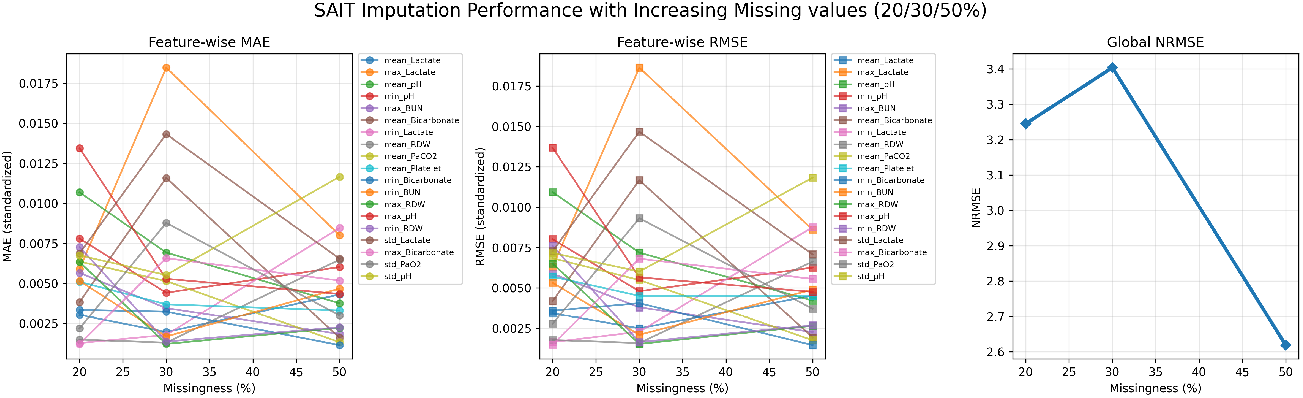
SAIT Imputation Performance with Increasing Missing Values

**Fig. 8:**
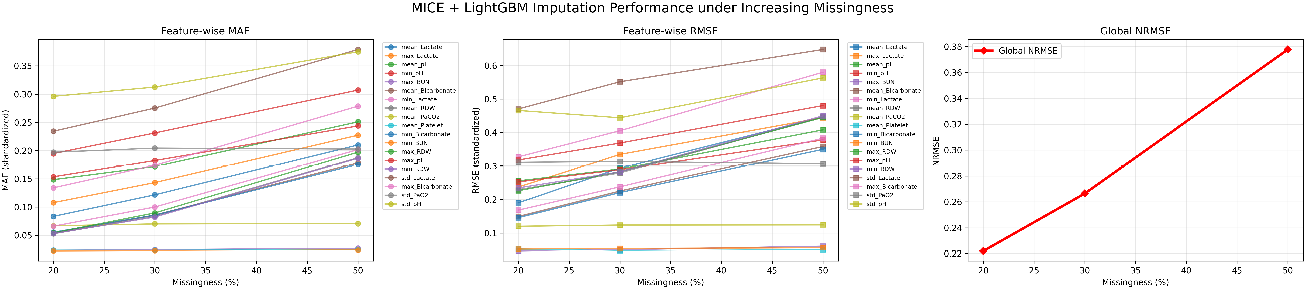
MICE+LightGBM Performance under Increasing Missing Values

#### Lactate measurements (mean, max, min, std)

DAE and SAITS achieved consistently low MAE (0.001-0.018 range) across all missingness levels. MICE+LightGBM struggled particularly with lactate variability (std_Lactate), with MAE exceeding 0.3 at 50% missingness. pH measurements (mean, min, max, std): Deep learning methods maintained MAE below 0.014 across all conditions. MICE+LightGBM showed moderate performance on mean and min pH but poor performance on max_pH and std_pH, suggesting difficulty capturing the full distribution of acid-base balance.

#### Renal function markers (BUN)

All methods performed relatively well on BUN measurements, with DAE and SAITS achieving MAE below 0.006 and MICE+LightGBM below 0.028 even at 50% missingness. The lower dimensionality and stronger univariate patterns in BUN may explain this relative ease of imputation.

#### Electrolytes (Bicarbonate)

DAE consistently excelled in measuring bicarbonate levels, with MAE consistently below 0.007 in all instances. SAITS also performed very well at the more extreme levels of missingness; however, MICE+LightGBM had a significant degradation of performance at 50% missingness (MAE ¿ 0.25).

#### Hematology (RDW, Platelet)

SAITS excelled at measuring RDW with an MAE ¡ 0.007 for all methods of calculating missingness. DAE performed competitively; however, MICE+LightGBM struggled with respect to RDW variability, with MAE ¿ 0.195 at 50% missingness.

#### Respiratory (PaCO2, PaO2)

Both SAITS and DAE performed admirably using both methods of estimating PaCO2 and PaO2, with slightly better results from the SAITS model for PaCO2 and DAE for PaO2 variability than the other models.

The three visualisations provided by SHAP show how SAITS made predictions and identify which physiological variables most affected the extent to which those predictions would change. The SHAP beeswarm plot visualises the globally aggregated feature importance, providing a high-level overview of relevant predictors and their positive or negative impact on predictions across all samples, as shown in Figure 10. All the predictors, the mean PaCO_2_, mean pH, maximum pH, maximum lactate, standard deviation of PaO_2_, and minimum pH contributed the most to the predictions made by the SAITS model. The horizontal spread of the modelling shows how much a predictor can alter the prediction, and the colour gradient shows the magnitude of each predictor. For several important features, their effects were consistent: higher-than-expected values for those physiologic characteristics increased the predicted risk, whereas lower-than-expected values decreased it. High lactate or extreme acid-base values are two examples of predictors that may indicate a much higher risk for the patient when used in the model. This evidence strongly supports the model’s ability to link clinical meaning to predictors of tissue hypoperfusion, metabolic acidosis, and respiratory failure. Conversely, other physiological predictors, such as RDW, platelet count, bicarbonate, and BUN, also contributed to the risk, but to a lesser extent, as they tended to cluster around near-zero SHAP values. In addition, some patients had very high SHAP values for RDW, suggesting that SHAP values may reflect strong predictive factors that are independent of sample size, consistent with an ICU population that is heterogeneous.

**Fig. 9:**
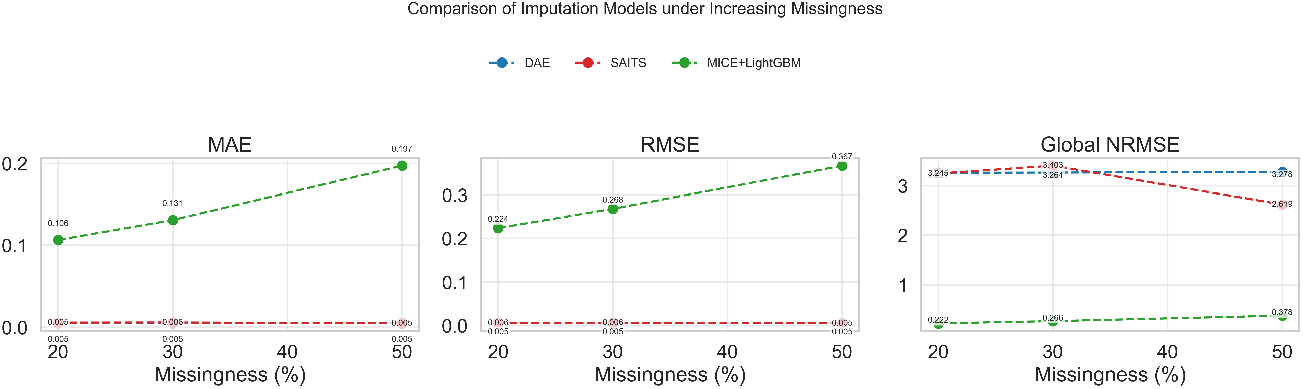
illustrates the trajectory of mean MAE, mean RMSE and global NRMSE with increasing missing values of DAE, SAIT and MICE+LightGBM

**Fig. 10:**
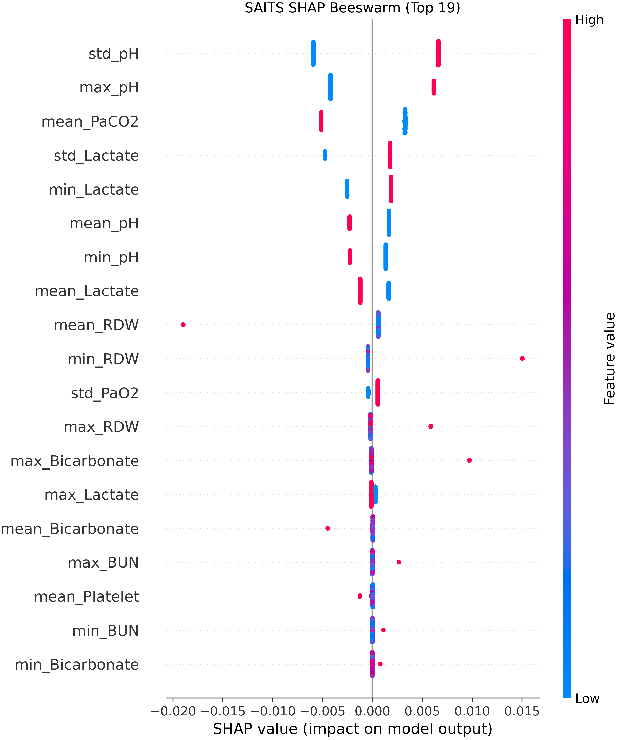
SHAP beeswarm plot for missing value imputation by the SAIT model

Figure 11 The SHAP decision plot shows how each prediction is made cumulatively using each of the predictors for each sample. In addition, very early on, there are some wide distances between models, making their predictions based on the inclusion of the highest-impact physiologic characteristics, particularly PaCO_2_, pH-related characteristics, and lactate, with all increasing distances after the addition of each of the other lower-ranked predictors. Eventually, the prediction curves converge towards each other and change much less dramatically, which means that the other predictors primarily fine-tune the prediction rather than drive it. The separation of the high-risk and low-risk problematic states from one another in a specified prediction path suggests that the model distinguishes between features based on biological meaning rather than random associations. Such behavior in clinical prediction models is desirable because it illustrates that the model’s predictions are a byproduct of meaningful biological patterns rather than the result of noise in the system.

**Fig. 11:**
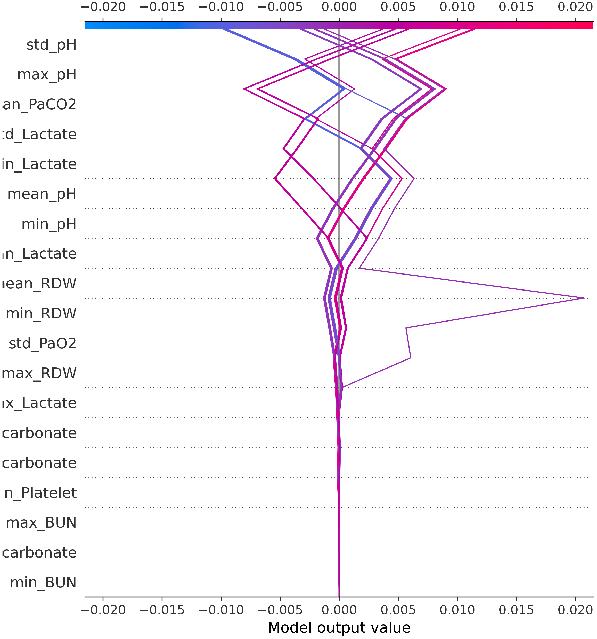
SHAP Decision plot for missing value imputation by the SAIT model

Figure 12 The SHAP violin plot further supports the effects of the features, when measured in terms of density of SHAP values associated with the predictor, for each predictor. Violin-shaped distributions of highly valued predictors, such as PaCO_2_, pH, and lactate, were wider and more separated based on their respective high and low feature values and effectively represented very strong and consistent predictive values, as seen in the results from the entire dataset. Additionally, features that were less important were shown to be more disposed to relatively narrow distributions with SHAP values centered around zero, and therefore contributed little globally to the prediction. The more symmetrical and structured distributions shown by the highly predictive features also indicate that there is a stable structure in the model. For example, the expected wider distributions of SHAP values for the lactate-related features indicate that there are more discriminative features present with lactates under different patient states than there would be for the other features. From a clinical perspective, the model’s description pattern correlates with the established physiologic predicator of mortality in the ICU. The predominant pathway to death for a patient in an ICU is related to ventilation markers (PaCO_2_), acid-base balance markers (pH and bicarbonate), perfusion markers (lactate), and the diversion from oxygenation markers (PaO_2_). Overall, these results suggest that the SAITS-imputed data maintained clinically valid relationships between the physiologic characteristics that were predictive for the patient population and showed a coherent physiologic pattern of prediction; therefore, they are both reliable for the imputation process and clinically interpretable for the prediction models made based on the imputed data.

**Fig. 12:**
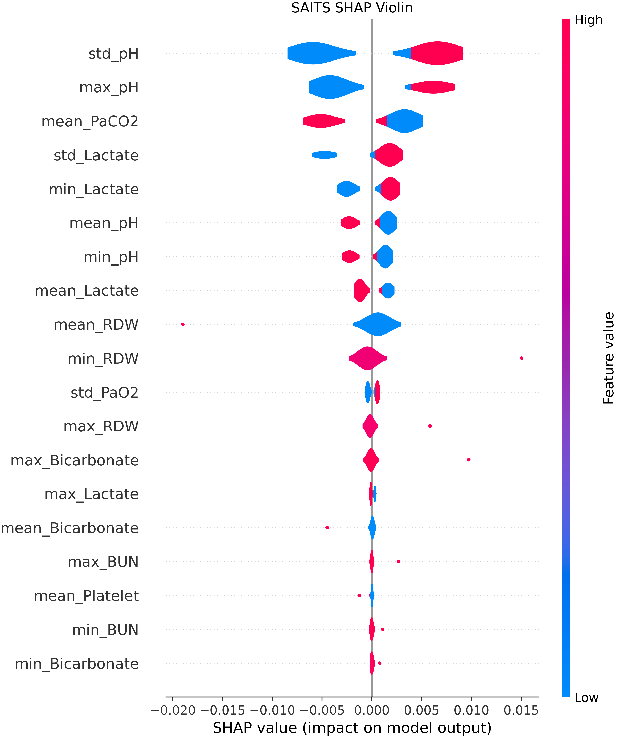
SHAP Violin plot for missing value imputation by the SAIT model

## 5 Discussion

### 5.1 Superior Performance of Deep Learning Methods

This study demonstrates that deep learning-based imputation methods—specifically Denoising Autoencoder (DAE) and Self-Attention Imputation for Time Series (SAITS)—substantially outperform the hybrid statistical-machine learning approach MICE+LightGBM for missing value imputation in heart failure ICU data. The performance advantage was consistent across all missingness levels (20%, 30%, 50%) and across diverse physiological feature types including lactate, pH, electrolytes, renal markers, and hematological parameters. The magnitude of improvement was striking: at 20% missingness, DAE achieved 34-fold lower MAE than MICE+LightGBM for mean_Lactate (0.002421 vs. 0.083502) and 19-fold lower MAE for mean_pH (0.00746 vs. 0.148502). These differences are not merely statistically significant but clinically meaningful, as imputation errors of the magnitude observed with MICE+LightGBM could lead to incorrect clinical inferences and suboptimal decision support. The superior performance of deep learning methods can be attributed to several architectural advantages. First, autoencoders and Transformer models learn joint distributions of all variables simultaneously through shared latent representations, enabling them to capture complex multivariate dependencies that sequential or univariate imputation methods miss [39][40]. Second, the nonlinear transformation capacity of deep neural networks allows modelling of intricate relationships between physiological variables that linear or tree-based models cannot represent [29]. Third, the end-to-end training of deep learning models optimises imputation directly for reconstruction accuracy, whereas MICE’s iterative procedure may accumulate errors across imputation cycles [41].

### 5.2 Robustness to Increasing Missingness

A critical finding is the differential robustness of methods to increasing levels of missingness. DAE and SAITS maintained stable performance as missingness increased from 20% to 50%, with NRMSE increasing by only 70% (from 3.260 to3.27). In contrast, MICE+LightGBM showed substantially greater degradation, with feature-averaged MAE increasing by 74% from 20% to 50% missingness. This degree of robustness is also important for clinical data, where the extent of missingness can vary considerably across patients, time frames, and clinical contexts. Consequently, if a method can perform well with minimal/low missingness but starts degrading in performance drastically with increasing levels of missingness, then it will have very little practical value in real-world applications. The consistent performance of the DAE/SADTS across missingness levels indicates that both models can learn robust representations of the underlying physiological data distribution that generalise well to situations with considerable information loss. In the case of SAITS, the attention mechanism may further contribute to the model’s ability to generalise across levels of missingness, as it uses dynamic weighting to prioritise variables and time points when utilising previously observed data patterns [42]. Thus, as the amount of missing data increases, the attention mechanism may allow the SAITS to efficiently focus on the most informative features during imputation, thus maintaining a high-quality estimate of the missing data. Similarly, the denoising objective in the DAE will also aid training for reconstructing data from heavily corrupted surfaces, thereby enhancing the ability of DAE/SAITS to perform successfully in a high-missingness environment [43].

### 5.3 Feature-Specific Performance Patterns

Feature-level analysis revealed interesting patterns in imputation difficulty and method-specific strengths. Standard deviation features (std_pH) were consistently the most challenging to impute, with higher MAE and NRMSE values across all methods. This is expected because these features represent temporal variability and are derived statistics rather than direct measurements, making them more abstract and potentially less correlated with other variables. MICE+LightGBM showed particularly poor performance on standard deviation features, with MAE exceeding 0.25 for std_Lactate at 50% missingness. This suggests that the iterative regression approach struggles to capture the distributional properties required to accurately impute variability measures. In contrast, the DAE and SAITS maintained the MAE below 0.015 for these features, indicating that their learned representations encoded information about the temporal dynamics and variability patterns. Renal function markers (BUN) were relatively easy to impute for all methods, likely because of their strong correlations with other clinical variables and more stable temporal patterns. Electrolyte measurements (bicarbonate) showed interesting method-specific patterns, with DAE demonstrating exceptional performance (MAE consistently below 0.007), whereas MICE+LightGBM struggled (MAE exceeding 0.35 at 50% missingness). This may reflect the complex relationship of bicarbonate with acid-base balance, respiratory function, and renal function, which can be captured by deep learning models through their multivariate representations.

### 5.4 Comparison with Literature

Our findings align with recent benchmarking studies on clinical time series imputation. [9] reported that SAITS, a Transformer-based model, achieved the best performance in most settings when compared to simple statistical methods, classical machine learning (MICE, MissForest), and other deep learning models (BRITS, US-GAN, GP-VAE) on MIMIC-IV monitoring data as shown in Table 8. Our results extend these findings to heart failure cohorts and aggregated tabular data derived from time series. The strong performance of denoising autoencoders is consistent with prior work showing that VAE-RNN combinations improve mortality prediction on MIMIC-III and MIMIC-IV through enhanced imputation [1]. While we did not evaluate VAE-RNN specifically, our DAE results suggest that autoencoder architectures generally provide robust imputation capabilities for clinical data. The poor relative performance of MICE+LightGBM contrasts with prior studies reporting strong performance of gradient-boosting methods for clinical data imputation [44][45]. However, those studies often evaluated imputation on datasets with lower rates of missingness or with different data structures. Our results suggest that while gradient boosting excels at supervised prediction tasks, its integration with MICE for imputation may not fully leverage its strengths, particularly under high missingness conditions.

**Table 8:**
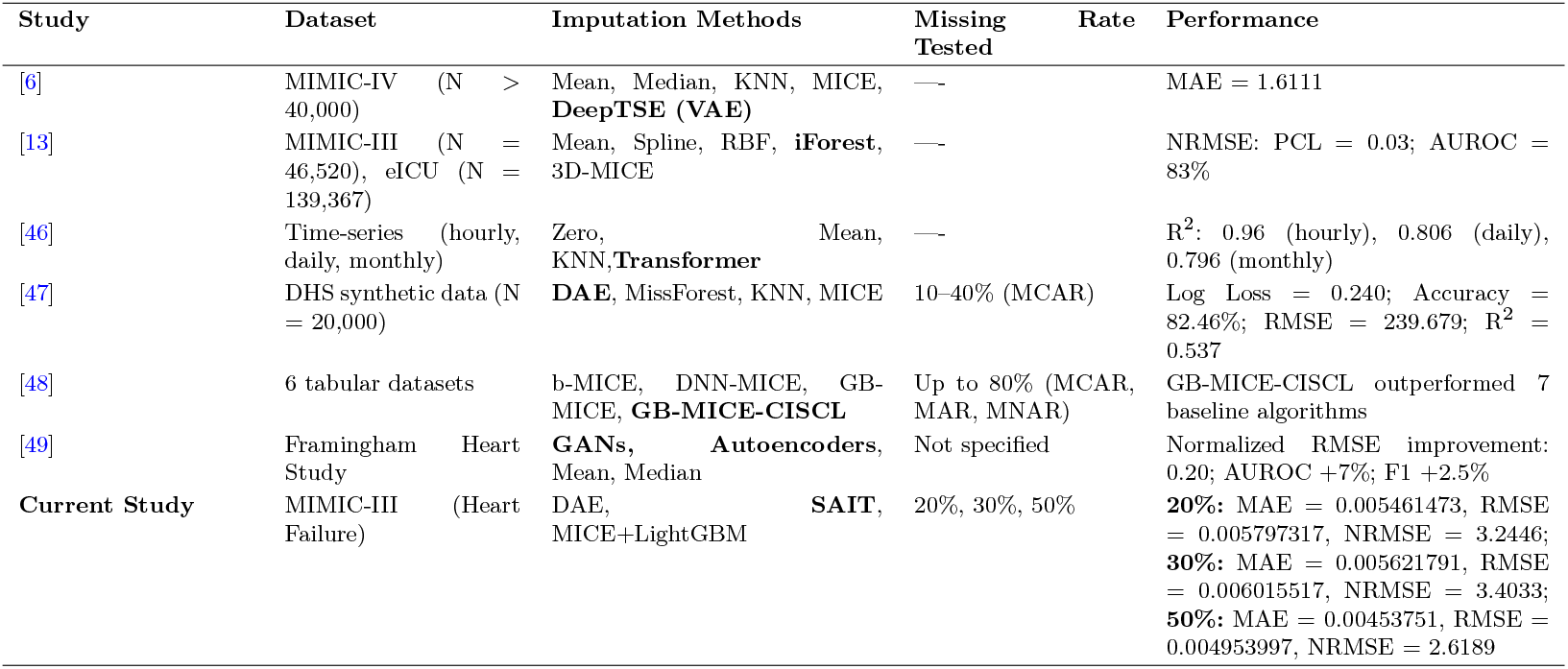
Comparison of Imputation Methods Across Clinical and Tabular Datasets.

### 5.5 Clinical Implications

Performance differences among these algorithms have critical implications for predicting heart failure severity in clinical decision support; therefore, understanding the underlying imputation errors, which can prevent reliable severity classification and lead to intervention errors in patients, is necessary to develop a clinically relevant system [50][51]. As lactate is a general marker of perfusion and shock, accurate determination of lactate level is essential for classifying patient risk and making clinical decisions [52]. The results suggest that the DAE and SAITS algorithms can predict mean_Lactate with a mean absolute error (MAE) of *<* 0.005 at 50% missingness; however, MICE + LightGBM achieves an MAE of 0.2104, which is almost 42 times greater. Such missing values may result in an inaccurate classification of the patient’s severity and subsequent inappropriate interventions. The same situation can be said for the measurement of pH and bicarbonate, both of which are critical for the assessment of acid-base balance in critically ill patient populations. [53] However, because deep learning algorithms can predict these types of values across the complete sample distribution (mean, min, max, and std), they can provide a more accurate picture of metabolic and respiratory dysfunction, thereby supporting a more rational clinical decision-making process. Moreover, the robustness of the DAE and SAITS algorithms allows for straightforward implementation and use in any clinical context, with minimal or no training required for each use. Finally, modern deep learning frameworks provide sufficient computational efficiency to support the use of these imputation algorithms in real-time clinical use [54].

Additionally, although this study explored interpretability using SHAP, future work should develop approaches that communicate model predictions in ways that are directly useful in clinical practice. Different stakeholders—including clinicians, patients, and policymakers—need customized explanations that align with their particular requirements and settings. Involving healthcare professionals in assessing the clinical relevance of the selected features is essential to ensure that data-driven findings are effectively translated into routine care.

## 6 Conclusion

According to this research, it was found that via Deep Learning (DL) based Imputation Methods (Denoising Autoencoder (DAE) and Self-Attention Imputation for Time Series (SAITS)) that MICE + Lightgbm performed poorly compared to DAE and SAITS on the imputation of missing values for heart failure patients using MIMIC-III. This outcome is consistent regardless of level of missingness (20%, 30%, 50%) as well as all physiological feature types (e.g. lactate, pH, electrolytes, renal markers, hematological). The MAE of DAE and SAITS was 10-100 times lower than that of MICE + Lightgbm across all features and was able to maintain stability with greater levels of Missingness, while MICE + Lightgbm showed significant levels of degradation over time with greater levels of Missingness. In addition to being superior to MICE + Lightgbm’s overall RMSE at all Missingness levels (particularly at severe levels of Missingness, i.e., 50%), both DL approaches also performed at a better level than MICE + Lightgbm for the imputation of temporal variability metrics. These results will have an important impact on the development of clinical decision support systems and predictive models.

Future research will concentrate on evaluating these approaches for realistic MAR and MNAR missing data, the performance of downstream applications of the data, raw time-series data, and prospective validation in real-life clinical settings. Additionally, developing uncertainty-aware deep learning imputation methods that produce both accurate point estimates and well-calibrated confidence limits could enhance their clinical utility. In summary, this study shows how much more powerful modern deep learning networks can be than traditional methods when it comes to imputing missing values for heart failure ICU records; hence, modern deep learning architectures should be integrated into clinical informatics programs and provide a basis for subsequent research in this important area of study.

## Data Availability

All data produced in the present work are contained in the manuscript

## Ethical Compliance

This study was conducted in accordance with internationally accepted ethical principles for biomedical research and complies with applicable data protection and privacy regulations. The research utilised fully de-identified secondary clinical data and did not involve direct interaction with human participants. Therefore, institutional ethical approval and individual informed consent were not required for this study.

The study adheres to ethical standards consistent with the Declaration of Helsinki and its subsequent amendments, as well as to comparable ethical frameworks governing retrospective analysis of anonymised healthcare datasets. All data processing, storage, and analysis procedures were performed on secure computing systems to ensure data confidentiality and integrity.

Special attention was given to the responsible use of artificial intelligence in healthcare. Model development and evaluation (SAITS, DAE, and MICE + LightGBM imputation frameworks) were conducted with transparency, reproducibility, and fairness considerations. Explainable AI (XAI) techniques, including SHAP-based interpretability and attention-based feature attribution, were implemented to improve model transparency and clinical interpretability.

## Data Access Statement

The dataset used in this study is the publicly available Medical Information Mart for Intensive Care (MIMIC-III) database. MIMIC-III is a large, de-identified clinical database containing detailed information on intensive care unit (ICU) patient admissions, including demographics, laboratory measurements, vital signs, medications, procedures, and clinical outcomes.

The dataset contains de-identified health-related data from approximately 60,000 ICU admissions and is widely used for machine learning and clinical prediction research. Researchers may request access after completing the required data-use training and certification. Official Access Link: https://physionet.org/content/mimiciii. The authors confirm that they complied with all data-use agreements and data-handling policies associated with the dataset.

## Author Contribution

Shilpa Sharma conceptualised the study, designed the methodology, performed data preprocessing, implemented the imputation and machine learning models, conducted experiments, analyzed the results, and drafted the manuscript. She also developed the SHAP-based interpretability analysis and prepared figures and tables. The co-author(s) Mandeep Kaur and Savita Gupta have contributed to study supervision, validation of methodology, critical revision of the manuscript, and provided domain expertise in clinical interpretation and research framing. All authors reviewed, edited, and approved the final manuscript.

